# Pediatrician and parental evaluation of child neurodevelopment at 2 years of age

**DOI:** 10.1101/2023.03.27.23287797

**Authors:** Giulia Segre, Antonio Clavenna, Elisa Roberti, Francesca Scarpellini, Massimo Cartabia, Chiara Pandolfini, Valeria Tessarollo, Ilaria Costantino, Maurizio Bonati

## Abstract

**Objective:** To assess the feasibility of a shared approach combining the clinical expertise of family pediatricians and parents’ perspectives in identifying potential disorders in children using standard clinical practice tools.

**Study design:** Within the NASCITA birth cohort, in addition to the family pediatrician’s clinical evaluation, the M-CHAT-R was completed by parents to assess the child’s language, social skills, behavior, and sensory areas. Parents were also asked to complete the PSI-SF to verify the magnitude of stress in the parent-child system. Univariate and multivariate analyses were performed to evaluate the association between child and parental characteristics and the presence of warning signs.

**Results:** The follow-up assessment was completed for 435 infants: 69 (15.8%) presented warning signs: 43 in the pediatrician’s assessment and 36 in the M-CHAT-R (10 in both). A total of 16 children (14 with warning signs) received a diagnosis after a specialist evaluation.

Being male (OR=2.42, 95%CI: 1.20-4.86) and having sleep disorders (OR=2.48, 95% CI 1.19-5.71) was associated with a greater likelihood of warning signs in the multivariate analysis, while reading aloud was a protective factor (not exposed versus exposed (OR=3.14; 95% CI 1.60-6.17).

For 73 children (18.4%), at least one parent tested positive for PSI-SF. An increased prevalence of parental distress was observed in children with warning signs (OR 2.36, 95% CI 1.27-4.37).

**Conclusions:** Integrating physician and parental perspectives during well-child visits and in clinical practice appears feasible and can improve the identification of children at risk of developmental disorders.

## Introduction

In the first few years of their lives, children develop the cognitive, social, and emotional skills that will provide the foundations for their lifelong health and achievements.^1^ An increased understanding of early neurobehavioral development is needed to prevent, identify, and treat childhood psychopathology effectively at the earliest possible stage^2^.

During early development, critical periods of brain plasticity are defined as those periods in which the development of brain functional properties is strongly dependent on and shaped by experience and environmental stimuli. Exposure to environmental stressors during critical periods of life can have negative long-term consequences for children’s health and development.^3^ Early interventions aim to prevent or minimize motor, cognitive, and emotional impairments in young children characterized by biological or environmental risk factors^4^ by acting in those first years of extremely plastic brain functioning.

Nowadays, there is an increase in formalized developmental screening in primary healthcare settings. Moreover, clinicians try to detect developmental disorders earlier because intervention services are more effective when provided in early childhood.^5^

The American Academy of Pediatrics^6^ recommends screenings at different age stages, based on different aims: during the first year of life of the child, paying particular attention to postpartum depression screening in mothers that can negatively affect child development^7^; at 9, 18, and 24 or 30 months of age, for the early identification of developmental disorders; and at 18 and 24 months, for the early identification of autism.^8,9^

In addition to promoting early identification of developmental and behavioral problems, an emphasis on parents as essential sources of information offers additional benefits. The role of parents as partners in the process of child health supervision is inherently emphasized, and the interest of the pediatric practitioner in the child’s overall functioning is highlighted.^10^

One of the factors that can have serious short- and long-term effects on children is the heightened stress experienced by their parents.^11,12^ Parental stress (maternal or paternal) is defined as the parent’s experience when the demands of the parenting role exceed the parent’s coping abilities.^13,14^ This perception can be caused by various social and environmental factors. Some research, for instance, showed effects on brain responses following postpartum depression, trauma, or substance abuse. In these scenarios, mothers present a reduced activation in response to infant cues, have lower sensitivity in, and gain less reward from, parenting, and are observed to be more anxious, intrusive, or traumatized.^15–18^

Current views emphasize a multifactorial and multi-determined conception of parenting stress involving both parent-related sources and individual distress.^19–21^ Within this framework, studies show that a dissatisfied relationship with the child is linked to less supportive reactions to the child’s negative emotions.^22^ Again, mothers’ negative emotionality correlates with unsupportive parental responses and subsequent child behavior problems.^23–25^

Crucially, evidence such as this suggests that a negative maternal experience represents a risk factor for developing behavioral problems and neuropsychiatric disorders in infancy. A recent study^26^ highlighted that parenting stress during infancy (11.4 ± 3.1 months of age) was significantly associated with mental health problems in 3-year-old children.

In this framework, as stated by the World Health Organization, identifying infants with warning signs indicating a heightened risk of a neurodevelopmental disorder is a crucial starting point for establishing a close relationship between parents and healthcare providers and setting up an early intervention.^27^ In particular, pediatricians are the first healthcare providers to strongly advocate for infants and children’s health and safety.^28–30^ They monitor development and are responsible for considering all aspects of a child’s well-being, including biological, social, and psychological factors.^30^

The aim of the present study was triple: (a) to assess the feasibility of using parent-reporting instruments to assess their perception of their children’s warning signs; (b) to ascertain whether there is an agreement between the family pediatricians’ (FP) clinical judgments of children’s warning signs and the parental perceptions; (c) to determine whether there is a correlation between parents’ distress and child development.

## Methods

### Participants and procedure

The Laboratory for Mother and Child Health of the Istituto di Ricerche Farmacologiche Mario Negri IRCCS in Milan set up the NASCITA Birth Cohort in collaboration with the National Paediatric Cultural Association (ACP).

The methods of the NASCITA study and the baseline cohort characteristics have been described elsewhere.^31,32^ Briefly, all Italian children receive primary health care exclusively from a family pediatrician until they are six years old as part of the national health system’s organization. The newborn population consists of all infants born during the enrolment period (April 1st, 2019– July 31st, 2020) and seen by the pediatricians for the well-child visits if parental consent was given. Seven well-child visits (within 45 days of life, at 3, 6, 12, 24, 36, and 72 months of age) are scheduled by the pediatrician in the first six years of a child’s life to monitor growth and development and offer preventive care.

The present study focuses on the assessment at two years of age: the questionnaires were completed by parents and FPs and collected during the well-child visit. Before starting with data collection, all pediatricians have been trained with online webinars to improve their competencies in completing the different tools.

We followed the Strengthening the Reporting of Observational Studies in Epidemiology (STROBE) reporting guideline.

### Measures

#### Pediatric assessment

Developmental milestones are commonly assessed to support pediatric surveillance and as part of developmental screening. For the pediatric assessment, the FPs, in addition to the routine questions on physical growth and health care check, fill in the CDC’s Learn the Signs, Act Early Milestones (LTSAE) Checklist at two years, which was adapted from the American Academy of Pediatrics’ Bright Futures Developmental Milestones checklist.

The CDC assesses the child’s language, cognitive/adaptive, motor, and social-emotional skills development (CDC-Developmental Milestones 2019, Bright Future 2020).^33^ This checklist is an evidence-informed developmental surveillance tool to identify early children with developmental delays and disabilities. The original CDC was translated from English into Italian. A back-translation was then performed by bilingual specialists who had never seen the original CDC. Three developmental and behavioral pediatricians then examined the Italian version. The back-translated versions were then compared with the original English version to assess the accuracy and resolve any discrepancies, as recommended by Jones, Lee, and Phillips (2001).^34^ To follow the Istituto Superiore di Sanità (Italian National Institute of Health – ISS) recommendations, four questions (included in the 18-24 months checklist) were added to the CDC checklist: 1) the child walks alone, kicks the ball or other object; 2) the child gets off the ground without support; 3) the child holds a pencil or a stick and scribbles on paper or the ground/floor; 4) when the child is denied something and/or has reactions of frustration, he can usually be calmed down quickly.

Moreover, two additional questions on the child’s development were added: the first one investigated the loss of competencies compared to the previous assessment, while the second one was related to the detection of hyperactivity signs by the FP.

FPs identified toddlers as “positive” for high risk if the total number of items that failed during the FP’s assessment was ≥ 8. This cutoff was defined by classifying scores in percentiles and considering the ones above the 95th percentile (i.e., a score of 8 or higher) as a warning sign.

### Parental assessment

The “Modified Checklist for Autism in Toddlers, Revised” (M-CHAT-R) is a measure developed to assess the risk for ASD in children ages 16-30 months. The M-CHAT-R is a simple screening tool considered valid for the early detection of warning signs in children compared to the conventional method (Italian translation: Salomone, Cecil, and Muratori, 2014).^35^ It is a checklist of 20 commonly observed child behaviors that requires the parent to report the presence of each specific behavior with a yes/no response. There are no culturally biased items. For all items except questions 2, 5, and 12, the response “NO” indicates a warning sign. The presence of abnormal behavior is assigned a score of 1, and the total score (obtained through the sum of all items where the response is identified as a warning sign) is interpreted. A total score of ≤2 indicates a low risk for ASD, and no further follow-up is recommended. When a total score between 3 and 7 is obtained, second-stage follow-up questions should be administered to decrease false-positive results: they were reassessed using M-CHAT-R Follow up questionnaire only on those items that originally failed. Children who were found to be in the low to medium-risk category were advised to have regular follow-ups. This assessment tool was chosen because some of the FPs involved in the study already used it as part of their routine clinical assessment. The strength of the M-CHAT-R is not only to detect children at risk of developing ASD but also to identify children that are likely to manifest a broader range of developmental disorders.

When a score of ≥8 is obtained, bypassing follow-up and immediately referring for diagnostic evaluation is allowed. For this study, children were considered as having ASD warning signs if they fell under the medium risk category (score ≥ 3).

To follow up on the child’s development, after nearly six or more months since their visit, we contacted the FP to ask whether the child had been referred for further evaluation to a neuropsychiatrist o psychologist and if a diagnosis was made.

### Parental distress

Data on parental distress were collected during the child’s two-year well-child visit. Thus, only a sub-cohort of parents voluntarily decided to fill in the questionnaire. Mothers and fathers were requested to complete the questionnaires separately; where this was not possible, we asked that at least one parent fill it in.

Parenting Stress Index - short form (PSI-SF),^13,36^ consists of 36 items measuring stress levels within the parent-child relationship. The respondent is asked to answer each item on a five-point Likert scale, ranging from 1 (strongly agree) to 5 (strongly disagree). The questionnaire yields a Total Stress score from three subscales. The Parental Distress subscale (PD; e.g., “I feel trapped by my responsibilities as a parent”) taps the parent’s perception of their behavior, including, for instance, perceived competence and sacrifices because of their parenting role. The Parent-Child Dysfunctional Interaction subscale (P-CDI; e.g., “My child rarely does things for me that make me feel good”) refers to the parent’s judgments on the interaction with their child. Finally, the Difficult Child subscale (DC; e.g., “My child makes more demands on me than most children”) measures the parent’s perception of his/her child’s personality, demandingness, hostility, and obedience. The Italian version of the PSI-SF^36^ has shown good internal consistencies ranging from α=.95 for the P-CDI subscale to α=.90 for the DC subscale. In the present study, for mothers, the internal consistency of the PSI-SF corresponds to α=.91 for the total stress scale. For fathers, the internal consistency of the PSI-SF corresponds to α=.92 for the total stress scale. A higher score suggests a higher stress level. Parenting stress percentile scores that fall between 15 and 84 are considered typical; scores above 85 indicate (at the 90th percentile) clinically significant parental stress. The total stress score (TS) is obtained by adding the scores of the three subscales PD, PCD-I, and DC. The test also includes a defensive response scale (DF) to check the validity of the protocol as it indicates whether the parent tends, for example, to give a better self-image or to minimize problems and perceived stress in the relationship with the child.

### Population profile covariates

Variables associated with a child presenting warning signs in previous studies were selected as covariates.^37–39^ The complete list of covariates (such as geographical area of residence, age of the parents at delivery, parental educational level, employment status, marital status and nation of birth (Italy yes/no), parity, pre-pregnancy BMI, gestational weight gain and chronic parental conditions) is reported in table 1. Mothers were grouped according to their pre-pregnancy BMI into three categories: underweight (≤18.5), normal (18.6-24.9), and overweight or obese (≥25.0). The weight variations recommended by the Institute of Medicine criteria were applied after grouping the mothers according to the pre-pregnancy BMI to evaluate gestational weight gain.^40,41^ We also considered whether delivery happened before or after the spread of the COVID-19 pandemic. Changes in birth assistance occurred, particularly during the first phase of the pandemic, with, for example, the impossibility of the fathers to enter the delivery room in many hospitals.^42^ In our sample, the pre-pandemic births happened between April 1st, 2019, and February 23rd, 2020, while the pandemic births occurred between February 24th, 2020, and July 31st, 2020.

**Table 1.** Association between child and parental characteristics associated with the presence of warning signs at parental or FP assessment

| <u>Variable</u> |  |  |  | <u>OR (95%CI)</u> | <u>p-value</u> |
| --- | --- | --- | --- | --- | --- |
| *Sociodemographic variables* |  |  |  |  |  |
|  |  | *Children at risk*<br>(N=69) | *Not at risk*<br>(N=366) |  |  |
| Geographical area of residence | *North* | 52 (75.4%) | 272 (74.3%) | 1.51<br>(0.68-3.33) | 0.29 |
|  | *Center* | 9 (13%) | 31 (8.5%) | 2.29<br>(0.80-6.50) |  |
|  | *South* | 8 (11.6%) | 63 (17.2%) | 1 |  |
| Both parents Italian | *Yes* | 56 (81.2%) | 330 (90.2%) | 1 | 0.03 * |
|  | *No* | 13 (18.8%) | 36 (9.8%) | 2.13<br>(1.06-4.26) |  |
| Maternal age at delivery | <30 | 13 (19.7%) | 83 (23 %) | 1 | 0.46 |
|  | 30-34 | 29 (43.9%) | 124 (34.3%) | 1.49<br>(0.73-3.04) |  |
|  | 35-39 | 16 (24.2%) | 112 (31%) | 0.91<br>(0.42-2) |  |
|  | >39 | 8 (12.1%) | 42 (11.6%) | 1.22<br>(0.47-3.16) |  |
| Paternal age at delivery | < 30 | 5 (7.6%) | 46 (12.8%) | 1 | 0.58 |
|  | 30-34 | 19 (28.8%) | 95 (26.5%) | 1.84<br>(0.65-5.24) |  |
|  | 35-39 | 21 (31.8%) | 121 (33.8%) | 1.60<br>(0.57 - 4.48) |  |
|  | >39 | 21 (31.8%) | 96 (26.8%) | 2.01<br>(0.71 - 5.68) |  |
| Maternal educational level** | *High* | 57 (82.6%) | 322 (88.2%) | 1 | 0.20 |
|  | *Low* | 12 (17.4%) | 43 (11.8%) | 1.58<br>(0.78-3.17) |  |
| Paternal educational level** | *High* | 48 (71.6%) | 295 (81.5%) | 1 | 0.06 |
|  | *Low* | 19 (28.4%) | 67 (18.5%) | 1.24<br>(0.96-3.16) |  |
| Maternal employment status | *Employed* | 48 (69.6%) | 280 (76.7%) | 1 | 0.21 |
|  | *Unemployed* | 21 (30.4%) | 85 (23.3%) | 1.44<br>( 0.22-2.54) |  |
| Marital status | *With partner* | 67 (97.1%) | 357 (97.8%) | 1 | 0.66 |
|  | *Single mother* | 2 (2.9%) | 8 (2.2%) | 1.33<br>(0.28-6.41) |  |
| *Parental risk factors:* |  |  |  |  |  |
| Maternal chronic conditions | *Yes* | 15 (21.7%) | 88 (24%) | 0.88<br>(0.47-1.63) | 0.68 |
|  | *No* | 54 (78.3%) | 278 (76%) | 1 |  |
| Paternal chronic conditions | *Yes* | 12 (17.4%) | 57 (15.6%) | 1.14<br>(0.57-2.25) | 0.71 |
|  | *No* | 57 (82.6%) | 308 (84.4%) | 1 |  |
| Pre-pregnancy BMI | *Underweight* | 4 (6%) | 32 (8.8%) | 0.75<br>(0.25-2.23) | 0.24 |
|  | *Normal* | 42 (62.7%) | 252 (69%) | 1 |  |
|  | *Overweight or obese* | 21 (31.3%) | 81(22.2%) | 1.56<br>(0.87-2.78) |  |
| Gestational weight gain | *Below* | 22 (32.8%) | 132 (36.5%) | 0.90<br>(0.49-1.65) | 0.77 |
|  | *Normal* | 28 (41.8%) | 151 (41.7%) | 1 |  |
|  | *Over* | 17 (25.4%) | 79 (21.8%) | 1.16<br>(0.6-2.25) |  |
| Physiological pregnancy | *Yes* | 52 (75.4%) | 308 (84.2%) | 1 | 0.08 |
|  | *No* | 17 (24.6%) | 58 (15.8%) | 1.74<br>(0.94-3.21) |  |
| Delivery during first pandemic wave | *Yes*<br>(24/02/2020 - 31/07/2020) | 9 (13%) | 54 (14.8%) | 0.87<br>(0.41 - 1.85) | 0.71 |
|  | *No*<br>(01/04/2019 - 23/02/2020) | 60 (87%) | 312 (85.2%) | 1 |  |
| *Child variables:* |  |  |  |  |  |
| Primiparous | *Yes* | 33 (47.8%) | 188 (51.5%) | 0.86<br>(0.52-1.44) | 0.57 |
|  | *No* | 36 (52.2%) | 177 (48.5%) | 1 |  |
| C-section delivery | *Yes* | 24 (34.8%) | 96 (26.2%) | 1.50<br>(0.87-2.59) | 0.14 |
|  | *No* | 45 (65.2%) | 270 (73.8%) | 1 |  |
| Healthy newborn | *Yes* | 57 (82.6%) | 312 (85.2%) | 1 | 0.58 |
|  | *No* | 12 (17.4%) | 54 (14.8%) | 1.22<br>(0.61- 2.42) |  |
| Newborn gender | *Male* | 46 (66.7%) | 186 (50.8%) | 1.94<br>(1.13-3.32) | 0.02* |
|  | *Female* | 23 (33.3%) | 180 (49.2%) | 1 |  |
| Skin to skin contact at birth | *Yes* | 48 (69.6%) | 281 (77.8%) | 1 | 0.14 |
|  | *No* | 21 (30.4%) | 80 (22.2%) | 1.54 (0.87-2.72) |  |
| Child sleeping disorders (from 6 months to 2 years) | *Yes* | 18 (26.1%) | 74 (20.2%) | 1.39 (0.77-2.52) | 0.27 |
|  | *No* | 51 (73.9%) | 292 (79.8%) | 1 |  |
| *Parental actions embedded in the nurturing care concept: prevention in pregnancy* |  |  |  |  |  |
| Mother smoker in pregnancy | *Yes* | 8 (11.6%) | 23 (6.3%) | 1.94 (0.83-4.55) | 0.13 |
|  | *No* | 61 (88.4%) | 341 (93.7%) | 1 |  |
| Mother consuming alcohol in pregnancy | *Yes* | 8 (11.6%) | 43 (11.9%) | 0.97 (0.43-2.17) | 0.94 |
|  | *No* | 61 (88.4%) | 318 (88.1%) | 1 |  |
| *Parental actions embedded in the nurturing care concept: prevention after birth* |  |  |  |  |  |
| Exclusive breastfeeding at 6 months | *Yes* | 22 (32.8%) | 119 (33.1%) | 1 | 0.96 |
|  | *No* | 45 (67.2%) | 240 (66.9%) | 1.01 (0.58-1.77) |  |
| *Parental actions embedded in the nurturing care concept: Approaches* |  |  |  |  |  |
| Reading aloud to children | *Yes* | 27 (39.1%) | 239 (65.3%) | 1 | < 0.0001 * |
|  | *No* | 42 (60.9%) | 127 (34.7%) | 2.93 (1.72-4.97) |  |
| Tummy time | *Yes* | 53 (76.8%) | 312 (86.2%) | 1 | 0.05 |
|  | *No* | 16 (23.2%) | 50 (13.8%) | 1.88 (1-3.55) |  |
| Bedtime routine | *Yes* | 13 (21.7%) | 87 (26.9%) | 1 | 0.39 |
|  | *No* | 47 (78.3%) | 236 (73.1 %) | 1.33 (0.69-2.58) |  |
| *Parental actions embedded in the nurturing care concept: Lifestyle* |  |  |  |  |  |
| Outdoor activities | *Yes* | 30 (44.1%) | 220 (62.1%) | 1 | < 0.01 * |
|  | *No* | 38 (55.9) | 134 (37.9%) | 2.08 (1.23-3.51) |  |
| Screen exposure | *Yes* | 16 (23.2%) | 97 (26.6%) | 1 | 0.55 |
|  | *No* | 53 (76.8%) | 267 (73.4%) | 1.20 ( 0.66-2.20) |  |
| Interacting with devices | *Yes* | 26 (40%) | 142 (39.7 %) | 1 | 0.96 |
|  | *No* | 39 (60%) | 216 (60.3%) | 0.99 (0.57-1.69) |  |
|  | *Yes* | 13 (21.7%) | 87 (26.9%) | 1 | 0.39 |
|  | *No* | 47 (78.3%) | 236 (73.1 %) | 1.33 |  |
|  |  |  |  | (0.69-2.58) |  |
| TV-on time in the home | Yes | 49 (74.2%) | 260 (76.5%) | 1 | 0.70 |
|  | No | 17 (25.8%) | 80 (23.5%) | 1.13<br>(0.62-2.07) |  |

\* *p-value of chi-square for trend test.* \*\*Educational level: low: no schooling or primary versus high: secondary school or university.

Concerning the perinatal and postnatal variables, type of delivery, skin-to-skin contact at birth, gender of the neonate, physiological development, and sleeping disorders were also analyzed. We also included the “healthy newborn” variable, which indicates those toddlers who were not born preterm, had no malformations at birth and were not admitted to the NICU in the postnatal period.

Moreover, different supportive parental actions embedded in the nurturing care concept and known to affect childhood health, development, and well-being positively were considered and were grouped into four general intervention areas, based loosely on concept and periods, as covariates. The areas were: 1) Prevention in pregnancy: no alcohol, no smoking during pregnancy; 2) Prevention after birth: exclusive breastfeeding at six months; 3) Approaches: Reading aloud, tummy time, bedtime routine; 4) Lifestyle: TV time in the home, screen time exposure, interacting with devices, outdoor activities (see Appendix 1 for a detailed description).

### Statistical analyses

Categorical variables were summarized using proportions and frequency distributions, and associations were tested using chi-square or Fisher’s exact test where applicable. Continuous variables were summarized using medians and interquartile ranges. Wilcoxon’s test was used to test differences in the distributions of continuous variables between two groups. Odds ratios (OR) were computed, and statistical significance was evaluated using 95% confidence intervals.

The sensitivity and specificity of pediatrician evaluation and M-CHAT-R in identifying children with a developmental disorder diagnosis were estimated.

Logistic stepwise regression analyses were used to identify variables associated with a greater likelihood of presenting warning signs at pediatrician or parental evaluation. Where data were missing, we used pairwise deletion so that all variables were used. All data management and analyses were performed using SAS software, version 9.4 (SAS, Institute Inc., Cary, NC, USA).

### Ethics

The study was approved by the Fondazione IRCCS Istituto Neurologico Carlo Besta’s Ethics Committee (February 6^th^, 2019, protocol n. 59).

## Results

A total of 48 FPs agreed to be involved in this sub-study, and 532 infants with their parents were recruited. Data concerning the follow-up assessment were available for 435 children (232 male and 203 female), which were included in the present study. No further information was collected for the other 97 toddlers due to the FPs’ retirement.

In all, 69 toddlers (15.9%) presented warning signs as either reported at the FP assessment or in the parental M-CHAT-R questionnaire. In particular, 33 children had warning signs only in the FP assessment, 26 only in the M-CHAT-R, and 10 in both parental and pediatrician assessments; 366 were not at risk.

Considering the children whose parents completed the M-CHAT-R, 399 (91.7%) were classified as low risk, 33 (7.6%) as medium risk, and 3 (0.7%) as high risk for autism spectrum or other neurodevelopmental disorders.

FP and parental assessments were mostly different, with only six overlapping questions (i.e., the ones evaluating pretend play, climbing onto furniture, pointing, walking, copying others, and following orders). For these, the agreement between parents and pediatricians was minimum, *k*=0.18 (95% CI -0.04-0.31).

For the 69 toddlers with warning signs, correlations were carried out with risk and protective factors to identify possible markers of risk conditions (Table 1). In the univariate analysis, being male (OR 1.94, 95% CI 1.13-3.32) or having at least one foreign-born parent (OR 2.13, 95% CI 1.06-4.26) were significant risk factors. On the other hand, protective factors were reading aloud (not exposed vs. exposed: OR 2.74, 95% CI 1.35-5.56) and spending at least one hour per day outdoors (not exposed vs. exposed: OR 2.08, 95% CI 1.23-3.51) (Table 1). The logistic regression analysis confirmed being male (OR 2.42, 95%CI: 1.20-4.86) as a factor associated with a greater likelihood of warning signs, in addition to having sleep disorders (OR 2.48, 95% CI 1.19-5.71), while reading aloud was confirmed as a protective factor, with children not exposed versus exposed having an OR=3.14 (95% CI 1.60-6.17).

Among the 69 toddlers with warning signs, 26 were referred for further evaluation to a neuropsychiatrist o psychologist.

In addition, 7 of the toddlers who did not present warning signs were also referred. Out of 33 children who underwent a specific clinical evaluation, 16 received a diagnosis (of whom 14 had warning signs and 2 did not): language disorders (n =9), autism (n = 3), developmental delays (n = 2), and other conditions (n = 2). For the M-CHAT-R, a sensitivity of 62.5% (95 % CI 35.4-84.8%) and a specificity of 76.5% (95 % CI 50.1-93.2%) were calculated. For the FPs’ assessments, a sensitivity of 75.0% (95% CI 47.6-92.7%) and a specificity of 41.2% (95 % CI 39.7-75.0%) were estimated. Combining the M-CHAT and FP assessments, we observed a sensitivity of 87.5% (61.7-98.4%) and a specificity of 29.4% (95 % CI 10.3-56.0%).

### Parental stress

At least one parent completed the PSI questionnaire for 397 children (61 with warning signs). Seventy-three children (18.4%) had one parent (mother or father) whose distress was considered clinically significant (≥ 85) in the total score. Nine children (5.4%) had both parents clinically distressed. The prevalence of parental distress was 14.9% among mothers (n= 368) and 13.8% among fathers (n = 195).

Among the 61 children considered at-risk by the parental or pediatrician assessments, 19 (31.1 %) had at least one parent whose distress was significant in the PSI. An association was found between the presence of warning signs in the children (either in the FP or parental assessment) and parental distress (OR 2.36, 95% CI 1.27-4.37). This association was confirmed with an increased prevalence of maternal (OR 2.42, 95% CI 1.23-4.76), but not paternal distress. Among the children referred for further evaluation, 9 (35 %) had at least one distressed parent, and among those who received a diagnosis, 4 (25%) had at least one distressed parent.

## Discussion

The first few years of life are a critical time in which the foundations of social and emotional competencies are laid. Monitoring neurobehavioral development is fundamental to preventing, identifying, and designing early treatment. While formalized developmental screening programs in primary healthcare settings are starting to be commonly used^9^, they often do not consider the shared perspectives of parents and pediatricians.

On the one hand, parents are the infants’ primary care environment. Factors such as parental stress and postpartum depression or trauma can result in reduced activation in response to infant cues, lower sensitivity, and less supportive reactions to the child’s emotions.^14,18,22^ These factors correlate with developmental risks of behavioral problems and neuropsychiatric disorders.^26^ For these reasons, parents may not always effectively ensure the best outcome for their children.^30^ Pediatricians are the first healthcare providers to advocate for infants’ health and safety^29^ and may have a crucial role in detecting early warning signs. Through clinical encounters with families and the knowledge of situations in the local community and the whole society, they can consider all aspects of a child’s well-being, including biological, social, and psychological factors.

The present study was the first to build a shared approach between parents and pediatricians in screening and encouraging early diagnoses.

In our cohort, 1 out of 6 children was considered “at risk” either at the FP assessment (10.1%), the parental M-CHAT-R questionnaire (8.3%), or both. Similar percentages were found in several international studies which used the CDC ^43^ or the M-CHAT-R questionnaire^44–47^. The combined risk percentage assessed in the present study was slightly higher, specifically 16%. The pediatricians, however, did not consider a psychiatric or psychological evaluation essential for most of these children. Because approximately 1 out of 20 children were referred for further evaluation, this proportion was, again, consistent with previous literature.^45^

For only 10 out of 69 children, the warning signs were reported by both parents and FPs. This number was expected since the tools (M-CHAT-R and CDC checklist) evaluate different skills and domains. It should be underlined, however, that when considering the six questions in the M-CHAT-R and the CDC questionnaires (evaluating pretend play, climbing onto furniture, pointing, walking, copying others, and following orders), the agreement between parents and pediatricians was low. This observation converges with the idea that parents and FPs may have different perceptions of the warning signs and that a shared approach is needed, as documented by the increase in sensitivity when combining the two evaluations.

Consistent with the findings of other studies,^48–50^ we have shown that signs of potential neurodevelopmental disorders are more frequent in males. Similarly, a male-female ratio of 2:1 exists among individuals with intellectual disability/developmental delay^51^ and a 4:1 ratio for individuals with autism diagnoses.^52^ As already demonstrated in other studies, we detected an association between sleep disorders in children and developmental disorders, such as pervasive developmental disorder (PDD) and attention deficit hyperactivity disorder (ADHD).^53,54^

Having at least one foreign parent was also associated with an increased likelihood of having warning signs. On the other hand, reading aloud and spending at least one hour per day outdoors were found to be protective factors: parents whose infants had a lower likelihood of warning signs were more adherent to those specific actions embedded in the nurturing care concept. Previous studies describe both these factors as having a protective role in neurodevelopment.^55,56^

From the parental assessment (PSI), we observed that almost one out of five children in our sample had one parent with clinically significant distress. The prevalence of parental distress was similar in mothers and fathers. In the literature, a lower prevalence of stress was reported by parents of a non-clinical sample, i.e., between 7 and 9%.^57,58^ Yet, it is important to highlight that no recent studies report updated estimates. Therefore, several factors (e.g., the COVID-19 pandemic, financial difficulties, and the environmental crisis) might have determined an increase in stress levels. Considering the children “at-risk,” this percentage rose to nearly one-third. Moreover, mothers and fathers of children and adolescents with neurodevelopmental disorders have reported high levels of stress in their parental role.^59^ An association was found between the presence of warning signs in infants and parental stress in mothers; the same was not confirmed in fathers. The same gender difference was previously reported when evaluating parents of children with cancer.^60^

Research has shown that the relationship between child behavioral problems and parental stress is bidirectional.^61^ We cannot exclude that parents with poorer psychological well-being may perceive and report their child’s behavior more negatively and may subsequently lack warmth and responsiveness.^62^ Nevertheless, the parents’ distress may also influence their childrearing, and the domestic environment may interfere with the parent’s ability to remain sensitive and accepting of the child.^63^ Children whose parents are significantly distressed might therefore receive less protective actions such as reading aloud and spending time outdoors. Awareness of such dynamics can guide FPs toward positively shaping parental behaviors and increasing protective activities. Moreover, it is also essential to consider that it is not possible to define whether parental stress is antecedent or consequent to the child’s difficulties or is an effect of the pandemic consequences.

Some limitations must be considered when interpreting this study’s results. The FPs involved participated voluntarily and might not be fully representative of all Italian family pediatricians. Secondly, in Italy, mothers are the caregivers who more frequently bring their children to the well-child visits; reduced paternal involvement in the present study confirmed this assumption. Greater paternal involvement in monitoring a child’s development should be encouraged. Even if neurodevelopmental disorders may be recognized at 18 months, the assessment was scheduled at 24 months consistently with the timing of well-child visits in most Italian regions. Therefore, in a few cases, the warning signs may have been present at an earlier time.

The COVID-19 pandemic represented a challenge for the NASCITA cohort study and may have impacted parents’ and children’s well-being. We included the period of birth (pre-pandemic vs. first wave) among the covariates, which did not influence the likelihood of developing warning signs or parental stress. Nonetheless, we cannot guarantee that the stressful situation that families had to face in the years after the children’s birth did not contribute to increasing parental stress and even affected child at-risk situations. Not all children with warning signs have been evaluated by a child psychiatrist or a psychologist, and therefore the number of children with a clinical diagnosis may be underestimated. On the other hand, it is likely that FPs decided to refer only children with a suspected developmental disorder, and in most cases, warning signs required only a follow-up by the FPs. Lastly, a diagnostic process might still be ongoing for some of the children. Therefore, it will be necessary to monitor the development of this cohort in future years to confirm the accuracy of the proposed assessment model.

Nonetheless, this study undoubtedly suggests practical interventions that FPs can advocate to support parents. Where significant stress is observed, parents might benefit from information on positive behaviors (i.e., supportive parental actions) to lower perceived stress and support a child’s development. Where this is not enough, FPs can recommend clinical follow-ups. Finally, this study demonstrated the feasibility of using the MCHAT as an assessment tool to detect various developmental disorders. Future studies might implement a similar approach asking parents to fill in different questionnaires to report on their child’s development.

## Conclusion

The present study demonstrated the feasibility and usefulness of child assessment that integrates the FPs’ and parents’ perspectives and evaluations. Thus, monitoring the infant’s development in several different contexts is crucial. Future directions should follow up with children and parents at later stages, possibly involving other figures with a primary role in children’s development, such as educators and teachers.

## Data Availability

All data produced in the present study are available upon reasonable request to the authors

## Abbreviations

NASCITA: (NAscere e creSCere in ITAlia);
M-CHAT-R: Modified Checklist for Autism in Toddlers, Revised;
PSI-SF: Parenting Stress Index, Short Form;
OR: Odds Ratio;
CI: Confidence Interval;
FP: Family Pediatricians;
LTSAE: Learn the Signs.
Act Early: ISS Italian National Institute of Health;
ASD: Autism Spectrum Disorder;
BMI: Body Mass Index;
COVID-19: Coronavirus Disease-19;
NICU: Neonatal Intensive Care Unit;
CDC: Centers for Disease Control and Prevention;
PDD: Pervasive Developmental Disorder;
ADHD: Attention-Deficit / Hyperactivity Disorder

## APPENDIX 1

*Parental actions embedded in the nurturing care concept*

Reading aloud to children: considered positive if a book had been read to the child in the two weeks prior to *each* of the three visits (6 months, 1 year and 2 year)

Tummy time: considered adequate if the child was placed belly down for any amount of time each day at 6 months. The American Academy of Pediatrics does not specify a precise amount of time a day, indicating a general 3-5 minutes, two to three times a day, increasing the time if the child enjoys it, for infants.^64^ The WHO guidelines recommend at least 30 minutes each day in a prone position while awake for 0-1 year-olds;^65^)

Bedtime routine: this was considered positive if there was a routine and it included a book or a song, and negative if it included tv/screen or “other”, at 1 and 2 year visit.^66,67^ We realize that classifying *“other” as negative is probably a bit stringent, but we considered the American Academy of Pediatrics’ suggestions for healthy sleep habits for children*.^66,68^

Screen exposure is considered positive if parents allow children to watch videos on screens (TVs, tablets, and smartphones) at 1 and 2 years.

TV-on time in the home: considered adequate if ≤4 hours per day at 1 and 2 year of age (Note: the reasoning behind this factor, and the cutoff time, was that, although a TV being on in the home will likely expose the child to a certain extent, the 4-hour cutoff allowed additional time for the parents’ watching of the news and of the TV when the children were in bed);^69^

The child was never given a smartphone or tablet to hold, and was never or almost never allowed to watch videos on any screen *at 1 year of age;*^70,71^

Interacting with devices: At both the 1 and 2 year visits, the parent was asked to indicate whether the child interacts with smartphone/tablet: (1 = never, 2 = sometimes, 3 = often). If the total score was 2 (never at both visits) the use was considered appropriate, if > 2 it was considered excessive.

Outdoor activities: At the 6 month, and 1 and 2 year visits, the parent was asked to quantify time spent by the child outdoors (1 = rarely, 2 = less than 1 hour per day, 3 = 1-3 hours per day, 4 = > 3 hours per day). If the sum of the scores was >8 it was considered adequate, if ≤ 8 it was considered insufficient.

Although no precise minimum amount of time outdoors to achieve health benefits in young children has been found, at least an hour a day at one year of age was considered adequate.^72–74^

## References

1 Shonkoff JP, Garner AS, The Committee On Psychosocial Aspects Of Child and Family Health, Committee On Early Childhood, Adoption, and Dependent Care, and Section On Developmental and Behavioral Pediatrics, Siegel BS, Dobbins MI, Earls MF et al. The Lifelong Effects of Early Childhood Adversity and Toxic Stress. Pediatrics 2012; 129: e232–e246.

2 Thompson L, Kemp J, Wilson P, Pritchett R, Minnis H, Toms-Whittle L et al. What have birth cohort studies asked about genetic, pre- and perinatal exposures and child and adolescent onset mental health outcomes? A systematic review. Eur Child Adolesc Psychiatry 2010; 19: 1–15.

3 Lautarescu A, Craig MC, Glover V. Prenatal stress: Effects on fetal and child brain development. In: International Review of Neurobiology. Elsevier, 2020, pp 17–40.

4 Guralnick MJ. Why Early Intervention Works: A Systems Perspective. Infants &Young Children 2011; 24: 6–28.

5 Rydz D, Srour M, Oskoui M, Marget N, Shiller M, Birnbaum R et al. Screening for Developmental Delay in the Setting of a Community Pediatric Clinic: A Prospective Assessment of Parent-Report Questionnaires. Pediatrics 2006; 118: e1178–e1186.

6 Foy JM, the American Academy of Pediatrics Task Force on Mental Health. Enhancing Pediatric Mental Health Care: Algorithms for Primary Care. Pediatrics 2010; 125: S109–S125.

7 Herba CM, Glover V, Ramchandani PG, Rondon MB. Maternal depression and mental health in early childhood: an examination of underlying mechanisms in low-income and middle-income countries. The Lancet Psychiatry 2016; 3: 983–992.

8 Catino E, Di Trani M, Giovannone F, Manti F, Nunziata L, Piccari F et al. Screening for Developmental Disorders in 3- and 4-Year-Old Italian Children: A Preliminary Study. Front Pediatr 2017; 5: 181.

9 Lipkin PH, Macias MM, COUNCIL ON CHILDREN WITH DISABILITIES, SECTION ON DEVELOPMENTAL and BEHAVIORAL PEDIATRICS, Norwood KW, Brei TJ, Davidson LF et al. Promoting Optimal Development: Identifying Infants and Young Children With Developmental Disorders Through Developmental Surveillance and Screening. Pediatrics 2020; 145: e20193449.

10 Glascoe FP, Dworkin PH. The Role of Parents in the Detection of Developmental and Behavioral Problems. Pediatrics 1995; 95: 829–836.

11 Crnic K, Low C. Everyday stresses and parenting. In: Handbook of parenting: Practical issues in parenting. Lawrence Erlbaum Associates Publishers., 2002, pp 243–267.

12 Deater-Deckard K, Panneton R. Unearthing the Developmental and Intergenerational Dynamics of Stress in Parent and Child Functioning. In: Deater-Deckard K, Panneton R (eds). Parental Stress and Early Child Development. Springer International Publishing: Cham, 2017, pp 1–11.

13 Abidin RR. Parenting stress index. Psychological Assessment Resources, Inc. (3rd ed.). Florida: Odessa., 1995.

14 Morgan J, Robinson D, Aldridge J. Parenting stress and externalizing child behaviour: Research Review: Parenting stress and externalizing child behaviour. Child &Family Social Work 2002; 7: 219–225.

15 Laurent HK, Ablow JC. The missing link: Mothers’ neural response to infant cry related to infant attachment behaviors. Infant Behavior and Development 2012; 35: 761–772.

16 Laurent HK, Ablow JC, Measelle J. Risky shifts: How the timing and course of mothers’ depressive symptoms across the perinatal period shape their own and infant’s stress response profiles. Dev Psychopathol 2011; 23: 521–538.

17 Musser ED, Ablow JC, Measelle JR. Predicting maternal sensitivity: The roles of postnatal depressive symptoms and parasympathetic dysregulation. Infant Ment Health J 2012; 33: 350–359.

18 Behrendt HF, Scharke W, Herpertz-Dahlmann B, Konrad K, Firk C. Like mother, like child? Maternal determinants of children’s early social-emotional development. Infant Ment Health J 2019; 40: 234– 247.

19 Moe V, von Soest T, Fredriksen E, Olafsen KS, Smith L. The Multiple Determinants of Maternal Parenting Stress 12 Months After Birth: The Contribution of Antenatal Attachment Style, Adverse Childhood Experiences, and Infant Temperament. Front Psychol 2018; 9: 1987.

20 Camisasca E, Miragoli S, Di Blasio P. Families with Distinct Levels of Marital Conflict and Child Adjustment: Which Role for Maternal and Paternal Stress? J Child Fam Stud 2016; 25: 733–745.

21 Lotto CR, Altafim ERP, Linhares MBM. Maternal History of Childhood Adversities and Later Negative Parenting: A Systematic Review. Trauma, Violence, &Abuse 2021; : 152483802110360.

22 Root AE, Rasmussen KE. Maternal Emotion Socialization: The Contribution of Inhibited Behaviour and Mothers’ Dissatisfaction with the Parent-Child Relationship: Brief Report. Inf Child Dev 2017; 26: e1955.

23 Eisenberg N, Fabes RA, Shepard SA, Guthrie IK, Murphy BC, Reiser M. Parental Reactions to Children’s Negative Emotions: Longitudinal Relations to Quality of Children’s Social Functioning. Child Development 1999; 70: 513–534.

24 Engle JM, McElwain NL. Parental Reactions to Toddlers’ Negative Emotions and Child Negative Emotionality as Correlates of Problem Behavior at the Age of Three: Parental Reactions to Toddlers’ Negative Emotions. Social Development 2011; 20: 251–271.

25 Dennis ML, Neece CL, Fenning RM. Investigating the Influence of Parenting Stress on Child Behavior Problems in Children with Developmental Delay: The Role of Parent-Child Relational Factors. Adv Neurodev Disord 2018; 2: 129–141.

26 Hattangadi N, Cost KT, Birken CS, Borkhoff CM, Maguire JL, Szatmari P et al. Parenting stress during infancy is a risk factor for mental health problems in 3-year-old children. BMC Public Health 2020; 20: 1726.

27 World Health Organization. Role of the Health Sector in Promoting Early Child Development. A Strategic Framework. 2011.https://apps.who.int/iris/handle/10665/205874.

28 Rudolf MCJ, Bundle A, Damman A, Garner M, Kaur V, Khan M et al. Exploring the scope for advocacy by paediatricians. Archives of Disease in Childhood 1999; 81: 515–518.

29 Wright CJ, Katcher ML. Pediatricians advocating for children: an annotated bibliography: Current Opinion in Pediatrics 2004; 16: 281–285.

30 Waterston T, Haroon S. Advocacy and the paediatrician. Paediatrics and Child Health 2008; 18: 213– 218.

31 Pansieri C, Pandolfini C, Clavenna A, Choonara I, Bonati M. An Inventory of European Birth Cohorts. IJERPH 2020; 17: 3071.

32 Pandolfini C, Clavenna A, Cartabia M, Campi R, Bonati M. National, longitudinal NASCITA birth cohort study to investigate the health of Italian children and potential influencing factors. BMJ Open 2022; 12: e063394.

33 CDC’s Developmental Milestones. 2019.https://www.cdc.gov/ncbddd/actearly/milestones/index.html.

34 Jones PS, Lee JW, Phillips LR, Zhang XE, Jaceldo KB. An Adaptation of Brislin???s Translation Model for Cross-cultural Research: Nursing Research 2001; 50: 300–304.

35 Robins DL, Barton M, Fein D. Modified Checklist for Autism in Toddlers, Revised with Follow-up. 2018. doi:10.1037/t67574-000.

36 Guarino A, Di Blasio P, D’Alessio M, Camisasca E, Serantoni M. Parenting Stress Index Short Form. Italian Validation. Giunti O.S. Organizzazioni Speciali.: Florence, 2008.

37 Carlsson T, Molander F, Taylor MJ, Jonsson U, Bölte S. Early environmental risk factors for neurodevelopmental disorders – a systematic review of twin and sibling studies. Dev Psychopathol 2021; 33: 1448–1495.

38 Kim SW, Youk T, Kim J. Maternal and Neonatal Risk Factors Affecting the Occurrence of Neurodevelopmental Disorders: A Population-Based Nationwide Study. Asia Pac J Public Health 2022; 34: 199–205.

39 Polańska K, Jurewicz J, Hanke W. Smoking and alcohol drinking during pregnancy as the risk factors for poor child neurodevelopment – A review of epidemiological studies. Int J Occup Med Environ Health 2015; 28: 419–443.

40 Nucci D, Chiavarini M, Duca E. Pre-pregnancy body mass index, gestational weight gain and adverse birth outcomes: some evidence from Italy. annali di igiene medicina preventiva e di comunnità 2018; : 140–152.

41 Benvenuti MB, Bø K, Draghi S, Tandoi E, Haakstad LA. The weight of motherhood: Identifying obesity, gestational weight gain and physical activity level of Italian pregnant women. Womens Health (Lond Engl) 2021; 17: 174550652110161.

42 Cesano N, D’Ambrosi F, Cetera G, Carbone I, Di Maso M, Ossola M et al. Maternity ward management and COVID-19 pandemic: Experience of a single center in Northern Italy during lockdown. Eur J Midwifery 2021; 5: 1–5.

43 Chorbadjian TN, Deavenport-Saman A, Higgins C, Chao SM, Yang JH, Koolwijk I et al. Maternal Depressive Symptoms and Developmental Delay at Age 2: A Diverse Population–Based Longitudinal Study. Matern Child Health J 2020; 24: 1267–1277.

44 Brennan L, Fein D, Como A, Rathwell IC, Chen C-M. Use of the Modified Checklist for Autism, Revised with Follow Up-Albanian to Screen for ASD in Albania. J Autism Dev Disord 2016; 46: 3392–3407.

45 Robins DL, Casagrande K, Barton M, Chen C-MA, Dumont-Mathieu T, Fein D. Validation of the Modified Checklist for Autism in Toddlers, Revised With Follow-up (M-CHAT-R/F). Pediatrics 2014; 133: 37–45.

46 Steinman KJ, Stone WL, Ibañez LV, Attar SM. Reducing Barriers to Autism Screening in Community Primary Care: A Pragmatic Trial Using Web-Based Screening. Academic Pediatrics 2022; 22: 263– 270.

47 Oner O, Munir KM. Modified Checklist for Autism in Toddlers Revised (MCHAT-R/F) in an Urban Metropolitan Sample of Young Children in Turkey. J Autism Dev Disord 2020; 50: 3312–3319.

48 Polyak A, Rosenfeld JA, Girirajan S. An assessment of sex bias in neurodevelopmental disorders. Genome Med 2015; 7: 94.

49 Thapar A, Cooper M, Rutter M. Neurodevelopmental disorders. The Lancet Psychiatry 2017; 4: 339– 346.

50 Fombonne E. Epidemiology of autistic disorder and other pervasive developmental disorders. J Clin Psychiatry 2005; 66 Suppl 10: 3–8.

51 Ropers HH. Genetics of intellectual disability. Current Opinion in Genetics &Development 2008; 18: 241–250.

52 Autism and Developmental Disabilities Monitoring Network Surveillance Year Principal Investigators, Centers for Disease Control and Prevention. Prevalence of autism spectrum disorder among children aged 8 years - autism and developmental disabilities monitoring network, 11 sites, United States. 2010.https://www.cdc.gov/mmwr/preview/mmwrhtml/ss6302a1.htm.

53 Matsuoka M, Nagamitsu S, Iwasaki M, Iemura A, Yamashita Y, Maeda M et al. High incidence of sleep problems in children with developmental disorders: Results of a questionnaire survey in a Japanese elementary school. Brain and Development 2014; 36: 35–44.

54 Hollway JA, Aman MG. Sleep correlates of pervasive developmental disorders: A review of the literature. Research in Developmental Disabilities 2011; 32: 1399–1421.

55 Weisleder A, Mazzuchelli DSR, Lopez AS, Neto WD, Cates CB, Gonçalves HA et al. Reading Aloud and Child Development: A Cluster-Randomized Trial in Brazil. Pediatrics 2018; 141: e20170723.

56 Scott S, Gray T, Charlton J, Millard S. The Impact of Time Spent in Natural Outdoor Spaces on Children’s Language, Communication and Social Skills: A Systematic Review Protocol. IJERPH 2022; 19: 12038.

57 Briggs-Gowan MJ, Carter AS, Skuban EM, Horwitz SM. Prevalence of Social-Emotional and Behavioral Problems in a Community Sample of 1- and 2-Year-Old Children. Journal of the American Academy of Child &Adolescent Psychiatry 2001; 40: 811–819.

58 Silva LMT, Schalock M. Autism Parenting Stress Index: Initial Psychometric Evidence. J Autism Dev Disord 2012; 42: 566–574.

59 Operto FF, Smirni D, Scuoppo C, Padovano C, Vivenzio V, Quatrosi G et al. Neuropsychological Profile, Emotional/Behavioral Problems, and Parental Stress in Children with Neurodevelopmental Disorders. Brain Sciences 2021; 11: 584.

60 Yeh C-H. Gender differences of parental distress in children with cancer. Journal of Advanced Nursing 2002; 38: 598–606.

61 Neece CL, Green SA, Baker BL. Parenting Stress and Child Behavior Problems: A Transactional Relationship Across Time. American Journal on Intellectual and Developmental Disabilities 2012; 117: 48–66.

62 Sanner CM, Neece CL. Parental Distress and Child Behavior Problems: Parenting Behaviors as Mediators. J Child Fam Stud 2018; 27: 591–601.

63 Guajardo NR, Snyder G, Petersen R. Relationships among parenting practices, parental stress, child behaviour, and children’s social-cognitive development. Inf Child Develop 2009; 18: 37–60.

64 Back to Sleep, Tummy to Play. HealthyChildren.org. https://www.healthychildren.org/English/ages-stages/baby/sleep/Pages/Back-to-Sleep-Tummy-to-Play.aspx (accessed 8 Jun2022).

65 World Health Organization. Guidelines on physical activity, sedentary behaviour and sleep for children under 5 years of age. World Health Organization: Geneva, 2019https://apps.who.int/iris/handle/10665/311664.

66 Toddler Bedtime Trouble: Tips for Parents. HealthyChildren.org. https://www.healthychildren.org/English/healthy-living/sleep/Pages/Bedtime-Trouble.aspx (accessed 8 Jun2022).

67 Janssen X, Martin A, Hughes AR, Hill CM, Kotronoulas G, Hesketh KR. Associations of screen time, sedentary time and physical activity with sleep in under 5s: A systematic review and meta-analysis. Sleep Med Rev 2020; 49: 101226.

68 Brush, Book, Bed: How to Structure Your Child’s Nighttime Routine. HealthyChildren.org. https://www.healthychildren.org/English/healthy-living/oral-health/Pages/Brush-Book-Bed.aspx (accessed 14 Jun2022).

69 Why to Avoid TV for Infants &Toddlers. HealthyChildren.org. https://www.healthychildren.org/English/family-life/Media/Pages/Why-to-Avoid-TV-Before-Age-2.aspx (accessed 8 Jun2022).

70 Madigan S, Browne D, Racine N, Mori C, Tough S. Association Between Screen Time and Children’s Performance on a Developmental Screening Test. JAMA Pediatr 2019; 173: 244.

71 Zhao J, Yu Z, Sun X, Wu S, Zhang J, Zhang D et al. Association Between Screen Time Trajectory and Early Childhood Development in Children in China. JAMA Pediatr 2022. doi:10.1001/jamapediatrics.2022.1630.

72 Playing Outside: Why It’s Important for Kids. HealthyChildren.org. https://www.healthychildren.org/English/family-life/power-of-play/Pages/playing-outside-why-its-important-for-kids.aspx (accessed 8 Jun2022).

73 CFOC Standards Database | National Resource Center. https://nrckids.org/CFOC/Database/3.1.3.1 (accessed 8 Jun2022).

74 Toffol G, Reali L. Il benessere psico-fisico dei bambini migliora se frequentano spazi verdi. Quaderni ACP 2018; 25: 1–3.

